# SCALP HIGH FREQUENCY OSCILLATION RATE DEPENDS ON SLEEP STAGE AND DECREASES WITH TIME SPENT IN SLEEP

**DOI:** 10.1101/2021.04.19.21255728

**Authors:** Dorottya Cserpan, Richard Rosch, Santo Pietro Lo Biundo, Johannes Sarnthein, Georgia Ramantani

**Affiliations:** Department of Neuropediatrics, University Children’s Hospital Zurich, Switzerland; Department of Neurosurgery, University Hospital Zurich, Switzerland; University of Zurich, Switzerland; Klinisches Neurozentrum Zürich, University Hospital Zurich, Switzerland; Children’s Research Centre, University Children’s Hospital Zurich, Switzerland

**Keywords:** pediatric focal epilepsy, scalp EEG, high frequency oscillations, HFO, sleep

## Abstract

High frequency oscillations (HFO) in scalp EEG are a new and promising epilepsy biomarker. HFO analysis is typically restricted to random and relatively brief sleep segments. However, considerable fluctuations of HFO rates have been observed over the recording nights, particularly in relation to sleep stages and cycles. Here, we identify the timing within the sleep period and the minimal data interval length that allow for sensitive and reproducible detection of scalp HFO. We selected 16 seizure-free whole-night scalp EEG recordings of children and adolescents with focal lesional epilepsy (median age 7.6 y, range 2.2-17.4 y). We used an automated and clinically validated HFO detector to determine HFO rates (80-250 Hz) in bipolar channels. To identify significant variability over different NREM sleep stages and over time spent in sleep, we modelled HFO rate as a Poisson process. We analysed the test-retest reliability to evaluate the reproducibility of HFO detection across recording intervals. Scalp HFO rates were higher in N3 than in N2 sleep and highest in the first sleep cycle, decreasing with time spent in sleep. In N3 sleep, the median reliability of HFO detection increased from 67% to 79% to 100% for 5-, 10-, and 15-min data intervals, improving significantly (*p*=0.004) from 5 to 10 min but not from 10 to 15 min. In this analysis of whole-night scalp EEG, we identified the first N3 sleep stage as the most sensitive time window for HFO rate detection. N3 data intervals of 10 min duration are required and sufficient for reliable measurements of HFO rates. Our study provides a robust and reliable framework for implementing scalp HFO as an EEG biomarker in pediatric epilepsy.

## INTRODUCTION

High frequency oscillations (HFO) in scalp EEG are a new and promising non-invasive epilepsy biomarker providing added prognostic value, particularly in the pediatric population (Boran *et al*., 2019; Ohuchi *et al*., 2019; Nariai *et al*., 2020; Tsuchiya *et al*., 2020; Cserpan *et al*., 2021). Beyond the initial use of HFO to delineate the epileptogenic zone in epilepsy surgery, HFO are currently investigated as potential biomarkers of epileptogenesis, seizure propensity, disease severity, and treatment response (Jacobs and Zijlmans, 2020; Gotman, 2021). The utility of scalp HFO as EEG-biomarker in pediatric epilepsy has been substantiated by recent studies corroborating the correlation of scalp HFO rates with (1) epileptogenesis after a first epileptic seizure, regardless of aetiology (Klotz *et al*., 2021), (2) seizure propensity in the presence of a predisposing condition such as centrotemporal spikes (Kramer *et al*., 2019) or tuberous sclerosis (Bernardo *et al*., 2018), (3) disease severity in focal lesional epilepsy (Boran *et al*., 2019) as well as in a wide range of pediatric epilepsy syndromes (Toda *et al*., 2015; van Klink *et al*., 2016; Ikemoto *et al*., 2018; Nariai *et al*., 2020); (4) treatment response following the administration of drugs or epilepsy surgery (Kobayashi *et al*., 2015; Boran *et al*., 2019). HFO analysis in patients with epilepsy is typically restricted to random and relatively brief time periods of mostly 5-30 min, even in those undergoing long-term EEG recordings (Zelmann *et al*., 2014). However, the question remains whether all available data, over several nights, should be utilised for analysis or whether carefully selected segments suffice for clinically meaningful results (Fedele *et al*., 2019). Data selection will have to balance the need for stable estimates of, e.g., HFO localisation patterns and rates that accurately reflect network properties (Gliske *et al*., 2018; Fedele *et al*., 2019; Chen *et al*., 2021) against the benefits of shorter segments making this approach more widely applicable, even in short standard EEGs. This makes the appropriate choice of the most suitable time windows and sample size to ensure data quality and representativity essential.

HFO analysis is routinely performed in sleep to avoid contamination by muscle artefacts (Zijlmans *et al*., 2017). However, considerable fluctuations of HFO rates have been observed across sleep stages and cycles (Staba *et al*., 2004; Bagshaw *et al*., 2009; Dümpelmann *et al*., 2015; von Ellenrieder *et al*., 2017; Gliske *et al*., 2018), analogous to the significant modifications in the rates of spikes, the standard EEG-biomarker of epilepsy. For spikes, a meta-analysis based on both scalp and invasive EEG revealed higher occurrence rates in NREM (N3) sleep compared to other vigilance states (Ng and Pavlova, 2013). Similarly, pathological HFO rates were shown to be highest during NREM sleep in invasive EEG studies in drug-resistant focal epilepsy (Staba *et al*., 2004; Bagshaw *et al*., 2009; von Ellenrieder *et al*., 2017; Al-Bakri *et al*., 2018) and, most importantly, pathological HFO during NREM sleep were shown to best localise the epileptogenic zone (Klimes *et al*., 2019). Furthermore, pathological HFO rates have recently been shown to decrease with accumulated time in sleep in invasive recordings (von Ellenrieder *et al*., 2017), pointing to data from the first sleep cycle as being most best suitable for analysis, allowing the most sensitive detection of HFO. However, it is unclear if findings from invasive EEG in mainly adult cohorts with drug-resistant focal epilepsy undergoing presurgical evaluation apply to scalp EEG recordings across childhood and adolescence and in a broader range of epilepsy syndromes. Despite being crucial for implementing HFO as a clinical tool, the relationship between sleep and scalp HFO characteristics in pediatric epilepsy remains insufficiently explored.

To address the hypotheses that scalp HFO rate is highest in the N3 sleep stage and decrease with accumulated time in sleep, we retrospectively analysed whole-night scalp EEG recordings of children and adolescents with focal lesional epilepsy, implementing a previously validated automated HFO detector (Fedele *et al*., 2016, 2017; Boran *et al*., 2019; Cserpan *et al*., 2021). While our previous study addressed the impact of patient age on scalp HFO (Cserpan *et al*., 2021), here we investigated changes in scalp HFO rate across different sleep stages and cycles to verify whether sleep-related factors explain the variability in HFO rate during sleep and determine optimal data selection strategies to identify HFO rate distributions across the scalp reliably.

## METHODS

### Patient recruitment

We recorded whole-night video-EEGs from 72 children and adolescents (< 18 y) with epilepsy at the University Children’s Hospital Zurich between January 2020 and January 2021. For the current study focusing on the effect of sleep homeostasis on scalp HFO rates, we included 16 patients that 1) were diagnosed with focal lesional epilepsy based on electroclinical correlations and imaging findings, 2) had a whole-night scalp EEG recording obtained at a high sampling frequency (>1000 Hz), and 3) had no seizures during the recording night. The clinical purpose of whole-night EEG included presurgical evaluation and treatment monitoring.

### Scalp EEG recording & data selection

Patients underwent whole-night video-EEG with 21 electrodes placed according to the international 10-20 system at a 1024 Hz sampling rate using the Micromed® EEG recording system (Mogliano Veneto, Treviso, Italy). Impedances were typically ≤ 1 kΩ. Sleep stages were marked by experienced neurophysiologists according to the American Academy of Sleep Medicine (AASM) (Berry *et al*., 2017). We selected only N2, and N3 sleep stages for further analysis since muscle activity and movement artefacts in wakefulness and REM sleep interfere with HFO detection, leading to increased false positives (Zelmann *et al*., 2014). We divided the selected data into 5-min intervals for further processing. Scalp EEG intervals with visible artefacts and channels with continuous interference based on visual inspection were excluded from further analysis.

HFO detection and analysis were performed blinded to clinical characteristics of the patients, and the results from HFO analysis were not considered for clinical decision making.

### Automated HFO detection

To capture the HFO activity with the highest possible spatial resolution given the data, we re-referenced to a bipolar montage using all combinations of neighbouring electrodes and thus obtained 52 bipolar channels.

Scalp HFO detection was conducted with a clinically validated, automated HFO detector (Fedele *et al*., 2016, 2017; Boran *et al*., 2019; Cserpan *et al*., 2021) that operates in three stages. Stage I determines a baseline amplitude threshold in time intervals based on the Stockwell entropy value in the ripple band (80-250 Hz). Events exceeding the threshold are marked as events of interest (EoI). In Stage II, the detector selects all EoI that exhibit a high-frequency peak isolated from low-frequency activity in the time-frequency space. In Stage III, the detector rejects all EoI with amplitude ≥40 µV or signal-to-noise ratio <4 or co-occurring in channels of the two hemispheres. There was no further visual validation of the events, rendering the algorithm fully automated.

We calculated the channel-wise HFO rate for each patient by dividing the number of detected HFO on each channel by the duration of the analysed EEG recording. For modelling purposes, we used the HFO count, i.e., the number of HFO events detected in each 5-min data interval, on the channel with the highest HFO rate during the recording night.

We controlled for the clinical plausibility of scalp HFO rate distributions by comparing the localisation of the channel with the highest HFO rate with the localisation of spikes in scalp EEG and focal lesions in MRI.

### Modelling approach

We hypothesized that scalp HFO rates were modulated by sleep homeostasis. To identify significant effects of NREM sleep stage (N2, N3), and time spent in sleep on scalp HFO rates, we modelled the HFO rate as a Poisson process based on the methodology previously applied for HFO analysis in invasive EEG (von Ellenrieder *et al*., 2017). We assumed that HFO events are not overlapping and that time intervals between consecutive events are statistically independent (Nagasawa *et al*., 2012; Nonoda *et al*., 2016; von Ellenrieder *et al*., 2017). Primary variables of interest affecting HFO rates were the NREM sleep stage (N2, N3) and the time spent in sleep, determined as the elapsed time expressed in hours from the first sleep stage until waking up in the morning. We further included the delta band and the sigma band activity, estimated as the root-mean-square value of the bandpass filtered signal in the 0.5-4 Hz and the 10-16 Hz band during each 5-min data interval. The delta band and sigma band activity were calculated for the scalp channels F3-C4, F4-C4 and averaged, then normalised to have zero mean and unit variance for each analysed sleep stage and patient.

We used different combinations of the explanatory variables to create Poisson process models (N=15) to estimate the mean HFO rate for every 5-min data interval as a function of these variables and thus to provide statistical evidence for their contribution to the fluctuation of HFO rates during whole-night sleep. We considered the mean HFO rate calculated over the total analysed period and relative variations of the mean HFO rate as determined by the variables in the model. We used the Akaike Information Criterion (AIC) for model comparison (Burnham and Anderson, 2002) since its value indicates both the goodness-of-fit and the complexity of the model. The respective coefficients and the AIC values for all 15 models are given in **Suppl. Table 1**. Two models can be considered significantly different with 95% probability when the difference in the respective AIC values exceeds 6 (Burnham and Anderson, 2002). To evaluate the performance of the Poisson process model, we used leave-one-out cross-validation using all but one patient from our cohort to estimate the model coefficients and testing the result on one patient, then repeating this procedure for all patients. To further validate our modelling of dynamic changes in HFO, we compared the predicted HFO counts in the 5-min data intervals, 1) as given by the Poisson process model when including all patient data while training (Poisson process - train), 2) when cross-validating the model on the left-out patient (Poisson process - test) model, with 3) the constant mean HFO rate model, according to the mean absolute error for each patient (**Suppl. Table 2, Suppl. Figure 1**).

**Table 1:** Patient characteristics and HFO properties. Patient characteristics include the etiological substrate of their focal epilepsy and the lobar localisation of the presumed epileptogenic zone based on the electroclinical correlations and MRI findings. HFO properties include mean HFO rates in events/min in all analysed data intervals for the channel with the highest HFO rates in total and separately for N2 and N3 sleep. y: years, f: female, m: male, nr: number, FCD: focal cortical dysplasia; nr: number; L: left; R: right

| ID | Sex | Aetiology | Localisation of the epileptogenic zone | Mean HFO rate in total (HFO/min) | Mean HFO rate in N2 (HFO/min) | Mean HFO rate in N3 (HFO/min) | Channel with the highest HFO rates |
| --- | --- | --- | --- | --- | --- | --- | --- |
| 1 | m | Postnatal stroke | L hemispheric | 3.8 | 3.2 | 4.6 | Fp1-Fz |
| 2 | m | FCD | R temporal | 3.1 | 2.1 | 5.0 | T6-P4 |
| 3 | f | FCD | R fronto-central | 2.4 | 2.6 | 2.1 | F4-Cz |
| 4 | m | FCD | L parietal | 2.4 | 2.5 | 1.7 | F3-C3 |
| 5 | m | FCD | L fronto-central | 1.8 | 1.6 | 2.3 | T5-C3 |
| 6 | m | Hypothalamic hamartoma | L frontal | 1.6 | 1.6 | 1.7 | Pz-O1 |
| 7 | m | Perinatal stroke | R hemispheric | 1.4 | 1.3 | 1.4 | Fp2-F8 |
| 8 | f | FCD | R frontal | 1.3 | 0.3 | 2.6 | P4-O2 |
| 9 | f | Rasmussen's encephalitis | L hemispheric | 0.6 | 0.3 | 0.9 | C3-Fz |
| 10 | m | FCD | L frontal | 0.5 | 0.6 | 0.3 | Fp1-F3 |
| 11 | m | FCD | L temporo-parieto-occipital | 0.4 | 0.4 | 0.5 | Fp1-Fp2 |
| 12 | m | Hippocampal sclerosis | L temporal | 0.3 | 0.2 | 0.3 | F8-T4 |
| 13 | m | Hemiconvulsion-hemiplegia-epilepsy syndrome | R hemispheric | 0.2 | 0.2 | 0.8 | Fp1-Fp2 |
| 14 | f | FCD | L frontal | 0.2 | 0.2 | 0.3 | Fp1-F3 |
| 15 | m | Diffuse glioma | R temporal | 0.2 | 0.2 | 0.2 | Fp1-Fz |
| 16 | m | Perinatal stroke | L hemispheric | 0.1 | 0.1 | 0.2 | T3-P3 |

**Table 2.** Estimated coefficients of relative HFO rate variation for the best model. The relative HFO rate variation is defined as follows: a change of X times in delta band activity is associated with a change in the HFO rate in N3 sleep of 0.232 X times. Both delta- and sigma band activity of the included N2 and N3 data intervals were normalised to zero mean and unit standard deviation. The mean HFO rate is higher in N3 than in N2 sleep and in the presence of delta- and sigma band activity, and it decreases with time spent in sleep by 5.0% and 2.1% per hour in N2 and N3 sleep.

|  | N2 | N3 |
| --- | --- | --- |
| NREM sleep stage | -16.4% | 30.6% |
| Time spent in sleep | -5.0% | -2.1% |
| Delta band activity | 9.8% | 23.2% |
| Sigma band activity | 7.1% | 9.7% |

**Figure 1.**
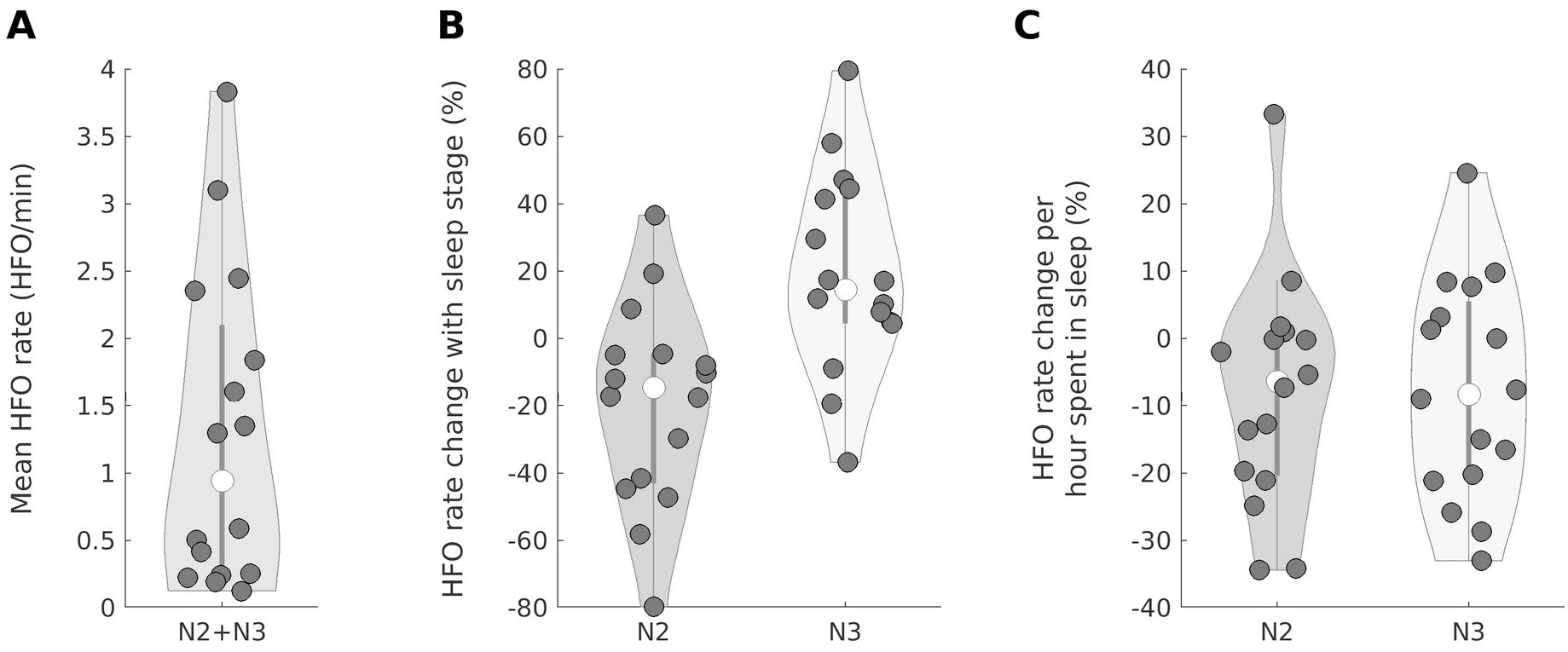
The mean HFO rate is higher in N3 and decreases with time spent in sleep. The violin plot shows the median (white circle) and the interquartile range (grey perpendicular line) of each distribution. A) HFO rate averaged over all N2 and N3 intervals for each patient. B) Compared to the mean HFO rate shown in panel A, the mean HFO rate is lower for N2 intervals and higher for N3 intervals. C) The Poisson process model indicates that the HFO rate decreases per hour spent in sleep (N2 median −6.3%, N3 median −8.3%).

### Test-retest reliability

To evaluate the reproducibility of scalp HFO detection, i.e. to investigate whether channel-wise HFO rates are consistent among different data intervals for each patient, we applied the test-retest reliability methodology as described in our previous work (Fedele *et al*., 2017). In short, we first calculated the normalised scalar products of HFO event vectors across different data intervals to depict the actual distribution and then created a null distribution from scalar products of HFO event vectors with permutated channel order. We report the mean number of scalar products with higher values than the 97.5 percentile of the null distribution, thus giving an estimation for the consistency of detected HFO rates in the analysed data intervals compared to randomised data. For this analysis step, we constructed 10-and 15-min data intervals for each patient by concatenating multiple 5-min data intervals.

### Statistics

We calculate the mean HFO rate over all recording intervals of each patient. Across patients, we describe distributions by their median and their interquartile range (iqr). To compare these distributions, we used non-parametric statistics. We used the Wilcoxon signed-rank tests to compare the reliability values between sleep stages and data intervals of 5-, 10-, and 15-min duration. To quantify correlations, we used Spearman’s rank correlation. Statistical significance was established at *p* < 0.05.

## RESULTS

### Patient characteristics, total length of sleep recordings, and HFO count

We included 16 patients (4 female) with focal lesional epilepsy (**Table 1)**. The median age at the time of the whole-night EEG recording was 7.6 y (range 2.2-17.4 y). Aetiology included focal cortical dysplasia in 8 cases, perinatal or childhood stroke in 3 cases, and single cases of diffuse glioma, hypothalamic hamartoma, Rasmussen’s encephalitis, hippocampal sclerosis, and hemiconvulsion-hemiplegia-epilepsy syndrome. The localisation of the presumed epileptogenic zone was frontal in 4 cases, temporal in 3, frontocentral in 2, parietal and temporoparietooccipital in one case each, and hemispheric in the remaining 5 cases.

We analysed 3605 min of EEG data, including 2500 min of N2 and 1105 min of N3 sleep. In total, we detected 4621 HFO: 2636 in N2 and 1985 in N3 sleep. The median length of analysed data per patient was 210 min (iqr 67.5), with a median of 237.5 (iqr 323) detected HFO per patient. The median length of analysed data per patient was 132.5 min (iqr 87.5) for N2 and 67.5 min (iqr 42.5) for N3 sleep.

### The scalp HFO rate is higher in N3 than in N2 sleep

The mean HFO rate over all data intervals (N2+N3) varied widely between patients (median 0.9 HFO/min, iqr 1.8), **Table 1, Fig. 1A**). The mean HFO rate was lower for N2 (median 0.5 HFO/min, iqr 1.6) and higher for N3 (median 1.2 HFO/min, iqr 1.9) (**Table 1**). Across all patients, mean HFO rates in N3 were higher than in N2 (median 29%, iqr 77; Wilcoxon signed-rank test, p= 0.049, z=1.96) (**Fig 1B**).

### The scalp HFO rate is higher in the first sleep cycle

Figure 2 presents the hypnogram of Patient 2 with the respective delta- and sigma band activity and the HFO rate during N2 and N3, including both the measured rate and the rate estimated by the Poisson process model. The figure illustrates the decrease of the HFO rate with time spent in sleep for Patient 2.

To analyse the HFO rate across all patients, we used the Poisson process model. Based on the AIC values (**Suppl. Table 1**), the best model was the one including all four variables, i.e., sleep stage, time spent in sleep, delta and sigma band activity. The coefficients of the relative HFO rate variation for all four included variables, as estimated by the best model, are given in **Table 2**. The mean HFO rate decreased with time with a relative median rate change of –6.3% (iqr 20.8) per hour in N2 and −8.3% (iqr 26.1) per hour in N3 sleep (**Figure 1C**).

**Figure 2.**
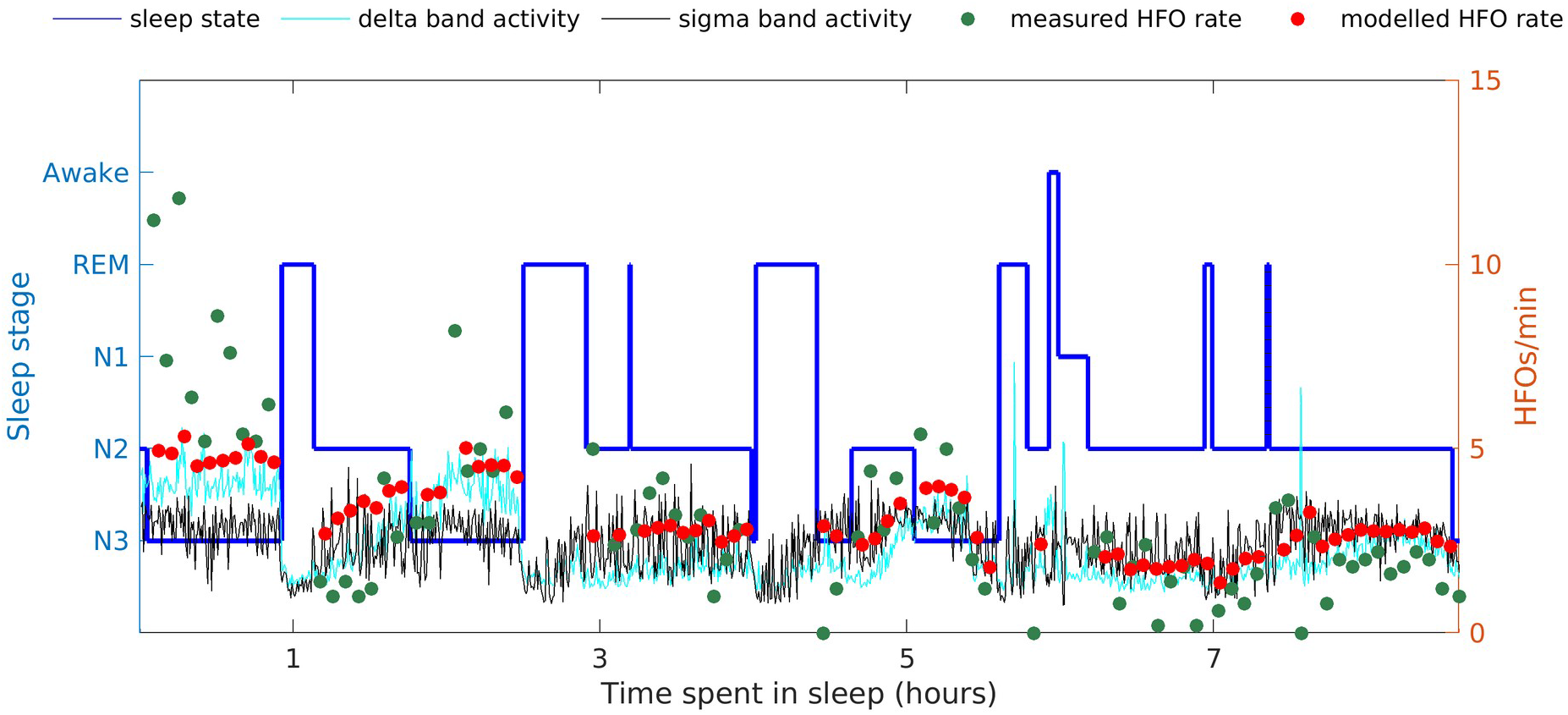
Hypnogram of a four-year-old patient with right temporal lobe epilepsy. We present the hypnogram (dark blue line), the delta- and sigma band activity (light blue and black line), the measured HFO rate (green dots) and the modelled HFO rate (red dots) in N2 and N3 sleep as a function of time spent in sleep. The measured HFO rate is highest in N3 sleep and decreases with time spent in sleep.

### Higher scalp HFO rates correlate with higher delta- and sigma band activity

Delta- and sigma band activity positively correlate with HFO rates during both N2 and N3 sleep stages, while this correlation is considerably stronger for delta-than for sigma band activity (**Table 2**). On a group level, an increase of one standard deviation in the delta band activity is associated with 23.2% higher mean HFO rates for N3 and 9.8% higher mean HFO rates for N2 sleep, whereas an increase of one standard deviation in the sigma band activity correlates with 9.7% higher mean HFO rates for N3 and 7.1% higher mean HFO rates for N2 sleep compared to the mean HFO rates.

### N3 data intervals of 10 min are required for HFO analysis

We calculated the test-retest reliability of HFO detection in 5-, 10-, and 15-min data intervals (**Fig. 3**). For 5-, 10-, and 15-min data intervals, the reliability of HFO detection was significantly higher in N3 than in N2 sleep (Wilcoxon signed-rank, 5-min: *p*=0.005, z=2.8; 10-min: *p*=0.013, z=2.5; 15-min: *p*=0.009, z=2.6).

**Figure 3.**
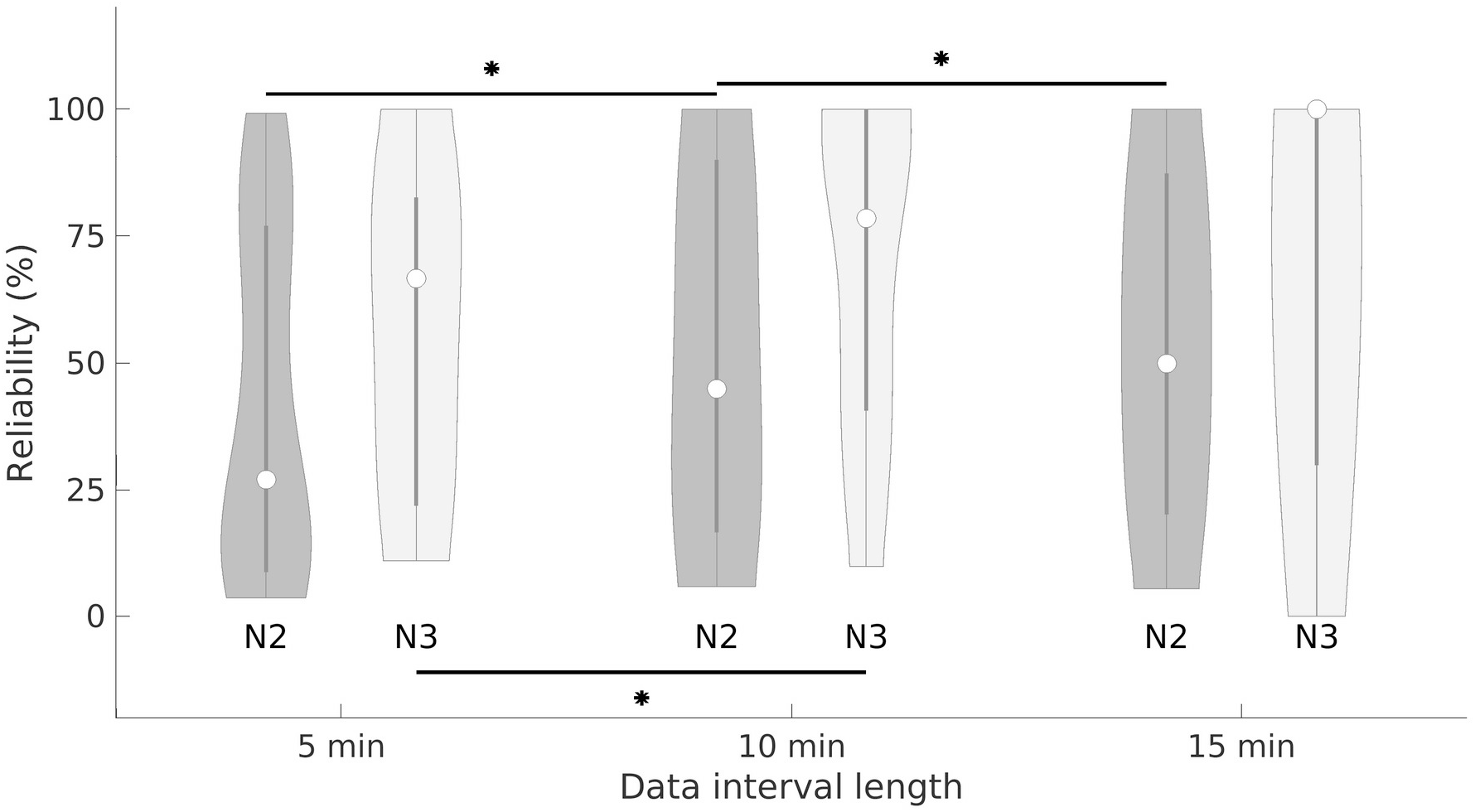
The test-retest reliability of HFO detection increases with the data interval length. The violin plot shows the median (white circle) and the interquartile range (gray perpendicular line) for each distribution. For N3 sleep, the median reliability rate of HFO detection increased from 67% (iqr 61) to 79% (iqr 59) to 100% (iqr 70%) for 5-, 10- and 15-min data intervals. The reliability increased significantly when increasing the analysed data interval length from 5 to 10 min (Wilcoxon signed-rank *p*=0.004, z=2.9). For N2 sleep, the median reliability rate of HFO detection increased from 27% (iqr 68) to 45% (iqr 73%) to 50% (iqr 67%) for 5-, 10- and 15-min data intervals. For 5-, 10-, and 15-min data intervals, the reliability of HFO detection remained significantly higher in N3 than in N2 sleep (Wilcoxon signed-rank, *p*=0.005, z=2.8, *p*=0.013, z= 2.5, *p*=0.009, z=2.6). \**p* < 0.05.

For N3 sleep, the median reliability of HFO detection increased from 67% (iqr 61) to 79% (iqr 59) to 100% (iqr 70%) for 5-, 10- and 15-min data intervals, with significant improvement in reliability when increasing the analysed data interval length from 5 to 10 min (Wilcoxon signed-rank, *p*=0.004, z=2.9), but not when increasing from 10 to 15 min (Wilcoxon signed-rank, *p*=0.953, z=0.1). For N2 sleep, the median reliability of HFO detection increased from 27% (iqr 68) to 45% (iqr 73%) to 50% (iqr 67%) for 5-, 10- and 15-min data intervals, with significant improvement in reliability when increasing the analysed data interval length from 5 to 10 min (Wilcoxon signed-rank, *p*<0.001, z=3.3) and from 10 to 15 min (Wilcoxon signed-rank, *p*=0.04, z=2.0).

Across all sleep stages (N2, N3) and data interval lengths (5-15 min), we established a strong positive correlation between the HFO rates and the test-retest reliability of HFO detection (Spearman’s rank correlation, *p*<0.05), suggesting that HFO detection is more reliable for patients with high HFO rates. However, it should be noted that longer data intervals ensure higher reliability values even for lower HFO rates. This observation suggests that longer intervals should be used for the later part of the night.

## 4. DISCUSSION

Electrophysiological markers of pathological epileptic brain activity show distinct dynamics associated with sleep stages and amount of sleep. This study demonstrates that scalp HFO in pediatric focal epilepsy change throughout whole-night sleep EEG recordings. To our knowledge, we are the first to determine the most sensitive time window in terms of sleep stage, cycle, and data interval length to ensure the quality and reproducibility of scalp HFO detection in pediatric epilepsy. We provide evidence that the first N3 sleep stage during a whole-night scalp EEG recording yields the highest HFO rates of the whole night recording. We demonstrate that reliable measures of HFO detection can be achieved in 10-min data intervals of N3 sleep, with higher HFO rates correlating with higher reliability values. Our observations enable selecting appropriate data intervals for stable HFO estimates in the first step towards their implementation as a valid epilepsy biomarker in a clinical setting.

### Scalp HFO rates are higher in N3 sleep

Scalp HFO rates were significantly higher in N3 sleep than N2 sleep in our study, in line with the significantly higher spike rates occurring in N3 sleep than other vigilance states (Ng and Pavlova, 2013). We may hypothesise that the higher scalp HFO and spike rates in N3 sleep are determined by the same neuronal processes that determine the decrease of spontaneous firing rates of cortical neurons with sleep (Vyazovskiy *et al*., 2009).

Moreover, the effects of NREM sleep stage on HFO rates in the scalp EEG of children with focal epilepsy reported here not only confirm previous observations deriving from the invasive EEG of adult patients (von Ellenrieder *et al*., 2017; Al-Bakri *et al*., 2018) but also extend these observations to a more accessible EEG modality and a much younger age group. Our study further demonstrates that including data from N3 sleep will increase the sensitivity of HFO detection because of the higher HFO rate in this sleep stage. This increased sensitivity is crucial for using HFO rate as a novel biomarker for epilepsy in the real-world clinical setting.

Finally, the remarkably high HFO rates in N3 sleep may be at least partly attributed to higher synchronicity levels in this sleep stage. This neuronal synchronisation results in the increased slope of slow-wave activity observed in N3 sleep and the amplitude of high-frequency activity. This state results in a higher phase-amplitude coupling between high (gamma, ripple) and low (theta or lower) frequencies in this sleep stage (Amiri *et al*., 2016). Our study replicated the strong positive covariation of HFO rate with delta band activity in N3 sleep, as previously suggested based on invasive EEG recordings (Nonoda *et al*., 2016; von Ellenrieder *et al*., 2017).

### Scalp HFO rate is higher in the first sleep cycle

The first N3 sleep stage during a whole-night scalp EEG recording in our study yielded the highest HFO rate, thus constituting the most sensitive time window for analysing HFO in pediatric epilepsy. Our findings are in line with the previously reported decrease of HFO rate with accumulated time in sleep in the invasive EEG of adults with focal epilepsy undergoing presurgical evaluation (von Ellenrieder *et al*., 2017), and confirm that the first sleep cycle is best suitable for studying HFO, irrespective of patient age and EEG modality.

The correlation of HFO rate with the accumulated time spent in sleep, in addition to their correlation with the different sleep stages, may be partly explained by the sleep-homeostatic changes in delta power showing a steady decline across the recording night (Riedner *et al*., 2007). This observation supports the notion that synchronisation, most pronounced during NREM slow-wave sleep, may be crucial for HFO generation (von Ellenrieder *et al*., 2017) and that sleep homeostatic changes in slow-wave amplitude along the night determine the HFO rate (Frauscher *et al*., 2015; Nonoda *et al*., 2016; von Ellenrieder *et al*., 2017).

In the animal model, the levels of glutamate in the cortical extrasynaptic space decrease during NREM sleep (Dash *et al*., 2009), whereas sleep deprivation leads to increased cortical excitability, resulting in a lowered threshold for epileptic activity (Badawy *et al*., 2006; Scalise *et al*., 2006). While slow-wave slopes are likely a function of neuronal synchrony, during late sleep, the periods of activity and inactivity of individual neurons became progressively less synchronised (Vyazovskiy *et al*., 2009).

### Reliability of scalp HFO detection

To confirm the reproducibility and establish the reliability of our scalp HFO detection, we performed a test-retest analysis, as previously developed by our group (Fedele *et al*., 2017), investigating the spatial profile of HFO rates across several EEG data intervals from each patient.

We showed that, while for higher HFO rates reasonably high reliability is reached even when using only 5-min data intervals, for lower HFO rates, longer data segments may prove indispensable. Nevertheless, it should be noted that the analysis of 10-min data intervals of N3 sleep provides considerably higher reliability than the analysis of shorter (5-min) data intervals.

Based on the findings from our cohort, we suggest that N3 data intervals of 10 consecutive minutes are required and sufficient for a consistent estimation of the spatial distribution of scalp HFO rate in most cases. We, therefore, recommend recording and analysing at least 10 min of N3 sleep to ensure a reliable scalp HFO detection in pediatric focal lesional epilepsy. HFO analysis requires stable spatial profiles over time that accurately reflect network properties since data quality and representativity will determine the validity of results (Fedele *et al*., 2019). It should, however, be noted that generalising these results to other datasets remains a hypothesis, especially for non-lesional/genetic epilepsy.

### Future directions

HFO have been shown to be modulated by sleep in all brain regions except for the frontal lobe in an invasive EEG study, including ten patients with frontal lobe coverage (Dümpelmann *et al*., 2015). To investigate the effect of lobar localisation of the HFO generator, larger cohort sizes than reported here are required.

Sleep stage but not sleep cycle has been demonstrated to determine the extent of HFO spread in an invasive EEG study focussing on the effect of sleep homeostasis on HFO (von Ellenrieder *et al*., 2017). Whilst standard EEG does not allow for evaluating such localised effects, similar questions may be investigated using high-density scalp EEG in the future (Fan *et al*., 2021). This outlook is especially relevant for the clinical setting, as high-density scalp EEG can be readily implemented in clinical epilepsy units (Zelmann *et al*., 2014).

## CONCLUSION

Our study provides a robust and reliable framework for implementing scalp HFO as an EEG biomarker in pediatric epilepsy. Based on our findings, restricting HFO analysis to 10-min data intervals of N3 sleep can increase the diagnostic yield while condensing the EEG recording time since these carefully selected segments should suffice for clinically meaningful results. This step would permit the application of scalp HFO in the screening of children at risk of developing epilepsy as biomarkers in the estimation of prognosis and question of treatment. Non-invasively detected scalp HFO may prove an essential resource for clinical assessment in a broad population of children affected by epilepsy.

## Data Availability

Further available data and code is indexed on our website https://hfozuri.ch

https://hfozuri.ch/resources/

## ACKNOWLEDGMENTS

We thank B. Alessandri, C. Carosio, L. Glaser, P. Hieber, and G. Selmin for their assistance with EEG recordings and data analysis. We thank the Swiss National Science Foundation (CRSK-3_190895 to G.R. and J.S.) and the Swiss League Against Epilepsy (Research Recognition Award to G.R.) for funding. The funders had no role in the design or analysis of the study.

## AUTHOR CONTRIBUTIONS

D.C. analysed data. D.C. and G.R. prepared figures and tables. R.R. and G.R. treated patients and monitored outcome. P.LB. acquired data. G.R. designed and supervised the study. D.C., J.S. and G.R. wrote the article. All authors critically reviewed the manuscript.

## PATIENT CONSENT

The collection of patient data and the scientific analysis were approved by and performed according to the guidelines and regulations of the local ethics committee (Kantonale Ethikkommission Zürich, KEK-ZH PB-2016-02055). All patients and their parents gave written informed consent before participating in the study.

## CODE AVAILABILITY

The software for the detection of HFO is freely available at the GitHub repository (https://github.com/ZurichNCH/Automatic-High-Frequency-Oscillation-Detector). Further available data and code is indexed on our website https://hfozuri.ch

## Abbreviation list

AASM: American Academy of Sleep Medicine
AIC: Akaike Information Criterion
DBA: Delta Band Activity
EoI: Events of Interest
FCD: Focal Cortical Dysplasia
HFO: High Frequency Oscillations
NREM: Non-Rapid Eye Movement sleep
SBA: Sigma Band Activity

