## Supplementary Material for "SCALP HIGH FREQUENCY OSCILLATION RATE DEPENDS ON SLEEP STAGE AND DECREASES WITH TIME SPENT IN SLEEP"

| Models | Variables | Coefficients | | AIC |
|---|---|---|---|---|
| | | N2 | N3 | |
| 1 | NREM sleep stage (%) | -16.64 | 30.61 | -1478.4 |
| 2 | Time spent in sleep (%/h) | -6.13 | -5.47 | -1347.2 |
| 3 | DBA (%) | 17.84 | 23.76 | -1475.8 |
| 4 | SBA (%) | 12.38 | 21.94 | -1373.4 |
| 5 | Time spent in sleep (%/h) | -5.64 | -1.68 | -1518.9 |
| | DBA (%) | 16.33 | 22.12 | |
| 6 | Time spent in sleep (%/h) | -6.78 | -4.15 | -1454.4 |
| | SBA (%) | 13.57 | 19.21 | |
| 7 | DBA (%) | 15.17 | 19.39 | -1486.4 |
| | SBA (%) | 5.89 | 7.43 | |
| 8 | NREM sleep stage (%) | -16.5 | 30.6 | -1559.6 |
| | Time spent in sleep (%/h) | -5.0 | -7.1 | |
| 9 | NREM sleep stage (%) | -16.4 | 30.6 | -1687.1 |
| | DBA (%) | 14.6 | 31.0 | |
| 10 | NREM sleep stage (%) | -16.8 | 30.7 | -1590.9 |
| | SBA (%) | 10.7 | 28.7 | |
| 11 | Time spent in sleep (%/h) | -6.04 | -1.62 | -1534.4 |
| | DBA (%) | 12.44 | 17.84 | |
| | SBA (%) | 7.76 | 7.36 | |
| 12 | NREM sleep stage (%) | -16.2 | 30.6 | -1728.4 |
| | Time spent in sleep (%/h) | -4.6 | -2.2 | |
| | DBA (%) | 13.4 | 28.9 | |
| 13 | NREM sleep stage (%) | -16.7 | 30.7 | -1669.5 |
| | Time spent in sleep (%/h) | -5.6 | -5.4 | |
| | SBA (%) | 11.6 | 25.1 | |
| 14 | NREM sleep stage (%) | -16.5 | 30.6 | -1699.5 |
| | DBA (%) | 12.1 | 25.3 | |
| | SBA (%) | 5.5 | 9.8 | |
| 15 | NREM sleep stage (%) | -16.4 | 30.6 | -1746.1 |
| | Time spent in sleep (%/h) | -5.0 | -2.1 | |
| | DBA (%) | 9.8 | 23.2 | |
| | SBA (%) | 7.1 | 9.7 | |

**Supplementary Table 1. Comparison of Poisson process models with different sets of variables.**
Coefficients and Akaike information criteria (AIC) values for all 15 models, each considering different sets of the four explanatory variables: NREM sleep stage (N2, N3), time spent in sleep, sigma-band, and delta-band activity. Coefficients are given in percentage per hour (%/h) for time spent in sleep and percentage per one standard deviation change for all other variables. Model 15 includes all four explanatory variables and has the lowest AIC value, indicating the best performing model.
DBA: delta band activity; SBA: sigma band activity; AIC: Akaike Information Criterion.

| Patient Nr. | Mean HFO rate (events/min) | $NMAE_{CV}$ | $NMAE_{Const}$ | $NMAE_{Poisson}$ |
|---|---|---|---|---|
| 1 | 3.83 | 0.219 | 0.173 | 0.189 |
| 2 | 3.10 | 0.123 | 0.154 | 0.103 |
| 3 | 2.45 | 0.280 | 0.255 | 0.256 |
| 4 | 2.35 | 0.302 | 0.229 | 0.293 |
| 5 | 1.84 | 0.168 | 0.187 | 0.162 |
| 6 | 1.60 | 0.179 | 0.217 | 0.175 |
| 7 | 1.35 | 0.283 | 0.310 | 0.276 |
| 8 | 1.30 | 0.130 | 0.154 | 0.122 |
| 9 | 0.59 | 0.211 | 0.240 | 0.210 |
| 10 | 0.51 | 0.147 | 0.147 | 0.146 |
| 11 | 0.42 | 0.268 | 0.256 | 0.266 |
| 12 | 0.25 | 0.241 | 0.235 | 0.240 |
| 13 | 0.24 | 0.251 | 0.290 | 0.250 |
| 14 | 0.22 | 0.153 | 0.168 | 0.152 |
| 15 | 0.20 | 0.194 | 0.191 | 0.194 |
| 16 | 0.13 | 0.160 | 0.178 | 0.160 |
| Median | | 0.202 | 0.204 | 0.191 |

**Supplementary Table 2. Comparison of Poisson process, constant rate, and cross-validation models.** Comparison of the predicted HFO rates, as given by the Poisson process model, the constant rate model, and the cross-validation model with the measured HFO counts, according to the mean absolute error for each patient. To compare the predicted HFO counts in the 5-min data intervals with the measured ones, we calculated the normalised mean absolute error (NMAE) as follows, with λ = HFO rate, and N = number of data intervals: NMAE=Σ|λmodel−λmeasured|/(N· max(λmeasured)−min(λmeasured)). The median value of the NMAE is the lowest for the Poisson process model, whereas the NMAE increases for the cross-validation model and is the highest for the constant model. $NMAE_{CV:}$ NMAE for the cross-validation model; $NMAE_{Const:}$ NMAE for the constant model; $NMAE_{Poisson:}$ NMAE for the Poisson process model.

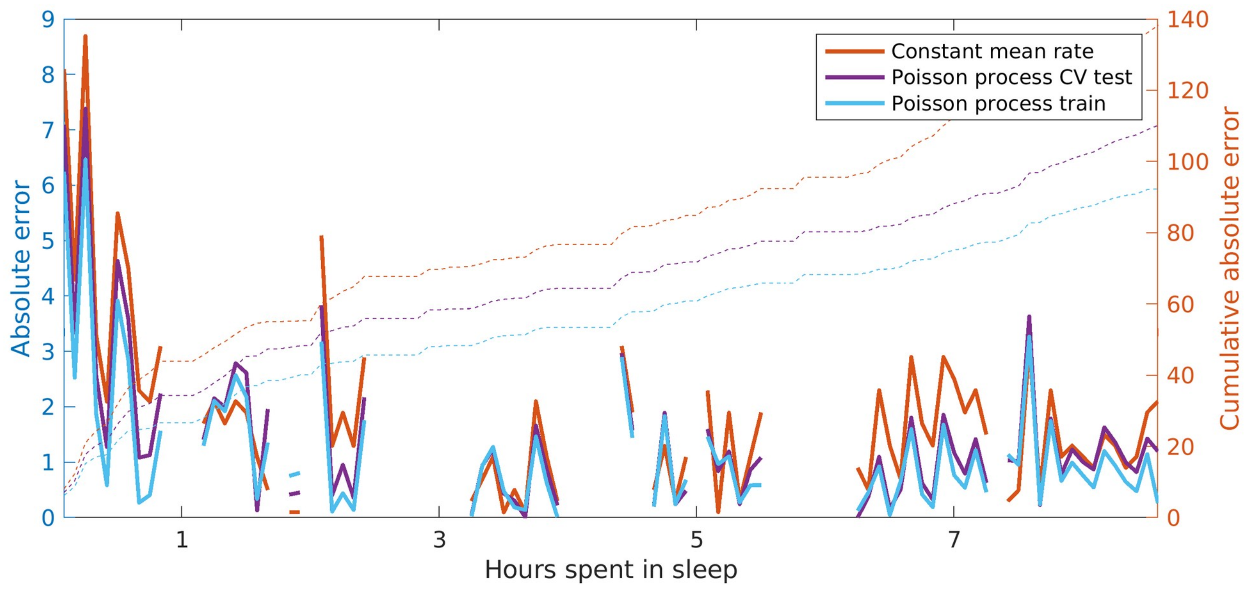

**Supplementary Figure 1: Comparison of Poisson process and the constant mean rate models.** Absolute error (line) and cumulative absolute error (dashed line) over time, as given by the constant mean rate model (orange) and the Poisson process model for Patient 2, when included in the training of the Poisson process model (blue) and when the data was only used for validation (purple). Over the recording night, as expected, the Poisson process model fitted on the full dataset performs better than the cross-validation setup, which in turn performs better than the constant rate model.
